# The impact of warning labels versus traffic light labels on food choice and negative emotional responses: two randomised controlled trials among UK adult populations with and without an eating disorder

**DOI:** 10.64898/2026.11.02.26367053

**Authors:** Rebecca Evans, Rebecca Latham, Amy Finlay, Eric Robinson

## Abstract

**Background:** In the UK, front-of-pack traffic light labels (TLL) are implemented voluntarily. The UK Government are considering alternative FOP approaches, such as nutrient warning labels (WL). However, the effects and potential harm of TLL versus WL in UK consumers are unclear. We compared the effects of WL, TLL, and no label on unhealthy food item selection and negative affect among UK adults with and without an eating disorder (ED).

**Methods:** We conducted two online randomised controlled trials. In Study 1, 924 UK adults without an ED diagnosis were randomly assigned (1:1:1) to a WL, TLL, or no labels condition and completed a food-choice task. In Study 2, 240 UK adults with an ED diagnosis completed a baseline no-label food choice task and were subsequently randomly assigned (1:1:1) to WL, TLL, or no labels for a follow-up food choice task. The primary outcome was the number of task-specific unhealthy food items selected. Negative affect before and after food choice tasks was measured as a secondary outcome.

**Findings:** In Study 1 (data collected 02/25 – 04/25), unhealthy food selection differed by condition (*p* = .013). Of five products selected, WL participants selected 0.20 fewer unhealthy items than TLL participants (8.0%; *p* = .038) and 0.28 fewer than controls (10.7%; *p* = .004). In Study 2 (data collected 11/25 – 02/26), unhealthy selection decreased by 0.39 items with WL (17.7%; *p* = .015), 0.13 with TLL (5.3%, p = .286), and 0.00 with no labels. The reduction was greater with WL than with no labels (*p* = .029). Label condition did not differentially affect task-related negative affect in either study (Study 1, *p* = .757; Study 2, *p* = .448).

**Interpretation:** WL tended to be associated with lower unhealthy food selection compared to TLL in UK adults. There was minimal evidence that WL or TLL caused negative emotional responses.

**Funding:** This research was funded by the National Institute for Health and Care Research (NIHR) Oxford Health BRC. The views expressed are those of the author(s) and not necessarily those of the NIHR or the Department of Health and Social Care.

**Registration:** ClinicalTrials.gov NCT07339969

## Introduction

Rising rates of obesity and non-communicable diseases (NCDs) are a significant public health concern. Approximately 63% of adults are living with obesity or classified as overweight and 1 in 5 children in England are leaving primary school with obesity (1). The likelihood of obesity is notably higher in areas of greater socioeconomic deprivation, where factors such as limited access to affordable healthy food and safe spaces for physical activity present a challenge (1). Tackling these issues requires multifaceted policy interventions.

Front of pack (FOP) labelling is a public health policy aimed at improving the diet of the UK population by providing comprehensible nutritional information on packaged food products (2). Systematic reviews and meta-analyses conclude that FOP labelling supports consumers to make healthier food choices (3,4). There is also evidence suggesting FOP labelling encourages reformulation of food products, but this varies by FOP label and implementation type (5).

In the UK, traffic light labels (TLL) have been implemented as a standardised voluntary FOP labelling system since 2013. TLL presents nutritional information per portion and uses green, amber, and red colour-coding to indicate if a food item is low, medium, or high in saturated fat, fat, salt, and/or sugar (6). Calorie content is also displayed but not colour-coded. Quasi-experimental evidence suggests that its introduction has improved the nutritional quality of household food purchases (7).

Chile introduced mandatory FOP nutrient warning labels (WL) on packaged foods high in calories, saturated fat, salt, and/or sugar in 2016 alongside other related policy interventions (8). Since then, several other counties have adopted or are considering adopting a FOP WL policy, including the UK, to help reduce rates of obesity and related NCDs (2,9). Research post-implementation of mandatory FOP WL has demonstrated a reduction in consumer purchase of high calorie, HFSS foods in addition to reformulation (10–12).

There is less research directly comparing effectiveness of TLL and WL. A systematic review and meta-analysis indicated that while both TLL and WL reduce the probability of selecting an unhealthy item relative to a control, WL perform better than TLL (4). It is theorised that this is due to WL being easier to process and emotionally respond to (e.g., concern about the high nutrient content). Whereas TLL may be more likely to confuse the participants if a product has, for example, both an amber and green label, which can lead participants to incorrectly perceive a product to be healthier than it is (13). The detailed numerical information and reference intake percentages displayed on TLL have also been highlighted as a point of confusion (14). However, this comparison was limited to eight studies with heterogenous label formats, position, study population, study design, and experimental settings (4). There are very few studies comparing TLL and WL directly in a representative UK sample using a realistic food choice task.

Understanding the relative costs and benefits of different FOP labelling systems is important to ensure optimal policy design and implementation. If labelling systems are well-understood and accepted, then they are more likely to be effective (15). Conversely, if individuals cannot understand the labelling system or feel annoyed by it, then this could lead to individuals ignoring labelling systems (16). It is also important to consider how vulnerable groups may respond to different FOP labelling systems. It is estimated that approximately 3.4 million or 1 in 20 people aged 16 or over in the UK have an eating disorder (ED) (17). People living with an ED are likely to be at increased risk of harm from food labelling. Most research to date into food labelling and individuals with an ED has centred on response to out-of-home (OOH) calorie labelling (18). A systematic review and meta-analysis found that ED pathology was related to being more aware of OOH labels, paying more attention to caloric intake, and adjusting behaviour in response to the labels (18). Qualitative findings suggest a polarised response to OOH calorie labels among individuals with an ED, ranging from them being reassuring and giving “a sense of control” to causing emotional distress and negatively impacting ED recovery (19). However, limited research has examined how different FOP labelling systems, including WL, may affect the behaviour of people living with an ED (20).

In a UK context, there is scarce research comparing FOP TLL to WL and no research has examined this in context of people living with an ED. Therefore, there is a need to (i) understand and compare how these different types of labelling systems influence food choice and (ii) identify any potential unintended psychological consequences, particularly among individuals with an ED.

The present research comprises two online RCTs in which participants made a series of hypothetical supermarket food choices across different food categories that either displayed WL, TLL, or no label (control). The aims of this work were to compare the impact of TLL and WL on food choice and negative emotional responses in (i) individuals without an ED (Study 1) and (ii) individuals with an ED (Study 2). Specifically, we hypothesised that both WL and TLL will affect how many unhealthy food items are selected (vs. control), but WL will have a larger impact on discouraging unhealthy food choices than TLL, and WL will have a larger impact on negative emotional responses than TLL.

We also examined calories and nutrients selected, and measures of participant perceived effectiveness and preference for the different FOP labelling systems.

## Methods

The studies were pre-registered on the Open Science Framework (https://osf.io/879kh) and ClinicalTrials.gov (NCT07339969). Both studies received institutional ethical approval (ref: 15576).

### Study design

In both studies, participants were recruited to take part in an online RCT where they were required to make hypothetical food choices under different FOPL conditions. In Study 1 (between-subjects), participants were randomly assigned (1:1:1) using the Qualtrics randomiser function to (1) WL group, (2) TLL group, or (3) no label group (control). In Study 2 (mixed design) participants completed a baseline food choice task in part one (no label present), before being randomly assigned (1:1:1) using the Qualtrics randomiser function to (1) WL group, (2) TLL group, or (3) no label group (in which they completed the same task a second time). The second part was required to be completed between 24 hours and 1 week after completing the baseline task. A mixed design was adopted for Study 2 to increase statistical power, because it was not feasible to recruit a similarly large sample as Study 1 when restricted only to participants with an ED.

### Participants

For Study 1, the inclusion criteria were current UK resident; over 18 years of age; fluent in English. The exclusion criteria were fasting or other form of restrictive eating e.g., intermittent fasting, elimination diet (FODMAP), religious restrictions, allergy restrictions, calorie/macronutrient restriction, food group restriction; clinical ED or disordered eating; dietary restrictions/intolerances including Vegan, Vegetarian, Gluten-free, Dairy-Free, Sugar-Free, Low salt. Recruitment was also stratified to be representative of the UK population in terms of sex, age, and education level (21–23), see Appendix A for specific stratification plan.

For Study 2, the inclusion and exclusion criteria were the same as for Study 1, except a current or past ED diagnosis was part of the *inclusion* criteria.

For both studies, participants were recruited using the online participant panel provider Prolific. For Study 2, Prolific recruitment was supplemented with targeted social media recruitment (posting in ED support groups on Facebook with permission) and the NIHR Be Part of Research registry. Be Part of Research is a free UK-wide service from the National Institute for Health and Care Research (NIHR), in collaboration with the NHS and the UK’s devolved administrations - with over 700,000 volunteers signed up. Individuals that were registered with Be Part of Research who expressed an interest in taking part in research relating to ‘eating disorders’ were identified by the service and sent information about the opportunity to take part in the study.

Prolific participants were paid at a rate of £6/hr. Participants recruited outside of Prolific were entered into a prize draw for a £100 Amazon voucher, and £1 was donated to the ED charity ‘Beat’ for each completed response.

### Procedures

Qualtrics XM survey software was used to create the survey questions and different trials within the food choice task. An online information sheet and consent form were automatically displayed to participants via Qualtrics before the survey started. Participants were required to confirm that they had read the information sheet, and consent to taking part, to proceed with the survey. The survey took approximately 15 minutes to complete. Upon completion of the survey, an online debrief sheet was displayed. In Study 2, an exit button was present on all pages which automatically directed participants to the debrief form, which included links to relevant support pages/phone numbers if the survey had triggered ED symptoms.

Each participant, irrespective of the group completed five trials within the main task. Participants viewed a selection of six packaged food items for scenarios involving labels predominately relating to (1) sugar (cereal), (2) salt (crisps), (3) fat (assorted savoury snacks), (4) saturated fat (pasta ready meals), and (5) calorie content (lunch salads) of the food products. Products were kept as similar as possible within each trial. For example, all cereals were chocolate flavoured and all crisps were salt and vinegar flavoured. Across the five scenarios, eleven products (36.7%) were not high in any nutrient, eleven (36.7%) were high in one nutrient, six (20.0%) were high in two nutrients, and two (6.7%) were high in three nutrients. See Appendix B for included items and ‘high in’ nutrient distribution per task. Pricing was not displayed to isolate the impact of the labels on food choice.

For each scenario, three items were ‘high in’ the key nutrient (e.g., sugar for the cereal scenario) and three items were not high in that nutrient. Criteria for products labelled ‘high in’ was consistent with labelling criteria for a red TLL in the UK; >25% of the nutrient reference intake per 100g, or >30% per portion (24). For the calorie content scenario only, >25% per 100g / portion (after rounding) was considered ‘high in’. The design of the WL was informed by the results from patient and public involvement (PPI) from a study co-designing and testing ‘high in’ WLs for food items in the UK (25). The design of TLL was as it exists on UK products currently, except calorie information was removed for all scenarios apart from the calorie-specific (lunch salads) scenario, in which calorie information was also colour-coded. Packaged products were selected from Tesco online supermarket. Label height was matched across TLL and WL conditions and was approximately consistent with TLL label size and placement on the Tesco website.

Participants were shown all six options at once and were asked to choose one item, imagining that they would actually pay for, receive, and eat this item. In total, participants chose five different items, one from each trial. After participants made their food choices, they were asked questions about the task they had just completed.

The tasks and questionnaires were identical in Study 1 and Study 2, unless indicated.

### Measures

### Participant characteristics

#### Sociodemographics

Participants were asked to report characteristics including age, sex, ethnicity, race, highest education level and household income. Education and household income were measured to characterise SEP. Self-reported weight and height were asked to calculate body mass index (BMI). Participants in Study 2 also reported the specific ED they have been diagnosed with.

#### Food choice motives

The Food Choice Questionnaire (FCQ) assesses the motives influencing an individual’s food choices (26). We included the health motives scale as health motivated individuals may be more likely to pay attention to and be influenced by a nutrition label when making food choices. The five items that measure health motives were included; mean score was calculated with a higher score indicating greater health motives.

#### The eating disorder examination-questionnaire short (EDE-QS)

The EDE-QS is a shorter version of the EDE-Q which provides a single overall score reflecting general ED symptom severity (27). This 12-item questionnaire with responses rated on a 4-point Likert scale, ranging from 0 (‘not at all’ / ‘0 days’) to 3 (‘markedly’ / ‘6-7 days’) was administered. The mean response to the 12 items was calculated. Higher scores indicate higher ED symptomatology.

### Primary outcome

#### Total number of unhealthy food items selected

Total number of selected unhealthy food items across the 5 trials (0–5). Unhealthy food items were coded as items eligible for a WL or a red TLL for the task-specific nutrient e.g., red for salt in the crisps trial.

### Secondary outcomes

#### Total calories and nutrients from selected food items

Total number of calories, salt (g), sugar (g), fat (g), saturated fat (g) from selected food items across the 5 trials was calculated.

#### Positive and negative affect schedule (PANAS)

Negative affect was assessed using the five PANAS items corresponding to negative affect (29). These items were included pre- and post- food choice task. Responses to ‘How afraid/upset/nervous/scared/distressed do you feel *right now*?’ were conveyed using a 5-point Likert scale ranging from 1 (‘Very slightly or not at all’) to 5 (‘Extremely’). Mean pre- and post- food choice task scores were calculated, with a higher score indicating greater negative affect.

### The below secondary outcomes were assessed in relation to the typical TLL (including calories) and WL (calorie WL included)

#### Negative arousal

An adapted four-item measure, previously used to assess responses to health WLs regarding snack selection (28) was included. Responses were conveyed using seven-point scales: ‘How afraid/worried/uncomfortable/disgusted does the label on food make you feel about consuming the product? With scores ranging from 1 (not at all afraid/worried/uncomfortable/disgusted) to 7 (very afraid/worried/uncomfortable/disgusted). A mean score was calculated for the four items, with a higher score indicating greater negative arousal.

#### Reactance and avoidance

Two items which have previously been used to measure response to health WLs on snacks were included (28); Reactance: ‘Are these labels annoying?’, Avoidance: ‘Are you likely to avoid these labels?’. Responses were examined using a 7-point Likert scale ranging from 1 (‘not annoying’ / ‘not at all’) to 7 (‘extremely annoying’ / ‘very likely’). Items were scored separately with a higher score indicating greater reactance / avoidance.

#### Acceptability of labels

Acceptability was assessed using the item ‘Do you support or oppose putting this label on food products?’. Responses were examined on a 7-point Likert scale ranging from 1 (‘strongly oppose’) to 7 (‘strongly support’). A higher score indicates greater support for the label.

#### Perceived message effectiveness (PME)

The three-item UNC PME scale was adapted for use (25,30). The three items were Discouragement: ‘This label discourages me from wanting to consume a food product’. Unpleasantness: ‘The label makes consuming a food product seem unpleasant’. Concern: ‘This label makes me concerned about the health effects of consuming a food product’. Responses were conveyed using a 5-point Likert scale ranging from 1(‘not at all’) to 5 (‘a great deal’). The mean response across the three items was calculated, with a higher score indicating higher PME.

### Additional outcomes

#### Label awareness

Participants were asked ‘Did you notice a nutrition label when making your food choices (e.g., when choosing crisps, pasta ready meal, cereal, savoury snacks, and lunch salad?’ (‘yes’ / ‘no’ / ‘unsure’).

#### Label influence

Participants were asked ‘Did the presence of a nutrition label on the food items influence your food choice?’ (‘Yes’ / ‘No’ / ‘Unsure’). If ‘yes’, how did the nutrition label influence your food choice? Response options (order of presentation randomised): ‘it made me select a healthier food item’ / ‘it made me avoid items high in fat/saturated fat/salt/sugar’ / ‘it made me select a less healthy food item’ / ‘it prompted me to think about my food choice before deciding’ / ‘it didn’t influence me and I ignored it’ / ‘not sure’.

#### Aim guessing

Participants were asked ‘What do you think the aim of this study is?’ with an open-text response option. Two researchers independently coded study aim awareness (‘aware’ / ‘unaware’) and resolved any disagreements through discussion. To be coded as ‘aware’, responses were required to mention the impact of labelling on food choice / emotion (or similar).

### Data analysis

#### Descriptives

Descriptive statistics were used to summarise participant characteristics, as well as primary, secondary, and additional outcomes. Continuous variables are summarised as means and standard deviations, and categorical variables are summarised as counts and percentages, split by labelling condition and overall.

### Study 1

Results from primary analysis were considered significant at p <.05. Results from all other analyses were considered significant at p <.01 to account for multiple analyses.

#### Primary analysis

A one-way ANOVA was used to assess the difference between the number of unhealthy food items selected in the three label conditions.

#### Secondary analyses

Separate one-way ANOVAs were used to assess the impact of the different label conditions on (i) total fat, saturated fat, sugar, salt, and calories in selected items, (ii) negative arousal (iii) reactance, (iv) avoidance, (v) acceptability and (vi) perceived message effectiveness.

A mixed ANOVA (within-subject factor: time point of negative affect rating, between-subjects factor: condition) was used to assess the impact of different label conditions on negative affect.

#### Exploratory analyses

Separate ANOVAs were used to assess interaction effects between label condition and relevant participant characteristics (age, sex, highest education, household income, health motives) in terms of (i) number of labelled items selected and (ii) negative affect.

#### Planned comparisons

For all primary, secondary, and exploratory analyses, planned follow-up t-test comparisons were used for any significant main effects or interactions (where relevant) to examine specific differences between the three label conditions.

#### Sensitivity analyses

Sensitivity analyses were conducted with aim guessers removed for primary and secondary analyses, to assess if results were the same after removing any participants that correctly guessed the study aims.

### Study 2

Analyses were the same as for Study 1, except mixed ANOVAs were used for outcomes measured at both baseline and follow-up, with time point as the within-subjects factor and label condition as the between-subjects factor. The condition x time interaction assessed whether change from baseline to follow-up differed between the three label conditions. For change in negative affect, a 3 x 2 x 2 mixed ANOVA was conducted, with condition as the between-subjects factor and time (pre-task or post-task) and session (baseline or follow-up) as within-subjects factors. For exploratory moderation analyses, interactions between condition, time and the relevant participant characteristic were examined. Outcomes assessed only at follow-up (e.g., negative arousal in response to the label) were analysed using between-subject ANOVAs. Baseline-adjusted ANCOVAs were conducted as sensitivity analyses for outcomes measured at both baseline and follow-up (see Appendix E).

#### Data exclusions

Participant data was excluded from relevant analyses if they (i) did not meet the inclusion criteria, (ii) failed both attention checks, and/or (iii) provided a data point deemed to be an outlier (> 2.5 the median absolute deviation [MAD] value from condition median) for primary or secondary outcomes (31). Participants were also excluded from Study 2 if they did not have usable follow-up data.

#### Power analysis

For both studies, we powered to our primary outcome (unhealthy food item selection). A meta-analysis found small effects of WL for reducing probability of selecting an unhealthy item relative to a control (OR = 0.65, or d = 0.24) and TLL (OR = 0.81, or d = 0.45) (4).

For Study 1, to detect a difference of d = 0.24 between conditions in follow-up independent samples t-tests (p < 0.05, 80% power, GPOWER 3.1) requires 274 participants per condition (total N = 822). We therefore aimed to recruit a total of 946 participants to account for loss of data (up to 15%).

For Study 2, to detect a mixed interaction effect of f = 0.12 (or d = 0.24) in our planned ANOVA (p < 0.05, 80% power, GPOWER 3.1), a total sample size of N = 246 is required. We aimed to recruit a minimum of 246 participants and up to a maximum of 400 participants (if feasible during the 3-month data collection period) due to uncertainty around effect size estimation for power calculation (as WL vs. TLL has not previously been tested in an ED population), and possible data exclusions.

## Results

A total of 968 participants completed Study 1. Participants were excluded if they failed both attention checks (n = 25) or reported an ED diagnosis (n = 19). This resulted in a final analytic sample of 924 participants.

A total of 403 participants completed Study 2. Participants were excluded for failing both attention checks (n = 8), not having a confirmed ED diagnosis (n = 66), or not having usable follow-up data (n = 79). This resulted in a final analytic sample of 240 participants. Most of the final sample were recruited through Prolific (n = 168, 70%) followed by social media (n = 58) 24.2%) and the Be Part of Research registry (n = 14, 5.8%).

Participant characteristics are presented in Table 1. For primary and secondary outcomes, outliers exceeding 2.5 MAD from the condition median were excluded from the relevant analysis (see Table 2 for final n for each outcome).

**Table 1:** Participant characteristics across both studies.

| Characteristic | **WL<br>(n = 306)** | **TLL<br>(n = 309)** | **Control<br>(n = 309)** | **Overall<br>(N = 924)** |
|---|---|---|---|---|
| **Study 1** |  |  |  |  |
| Age, years, M (SD) | 44.36 (12.97) | 45.17 (13.73) | 44.87 (13.59) | 44.80 (13.43) |
| Female, n (%) | 155 (50.7) | 162 (52.4) | 150 (48.5) | 467 (50.5) |
| Bachelor’s degree or above, n (%) | 154 (50.3) | 155 (50.2) | 149 (48.2) | 458 (49.6) |
| White ethnicity, n (%) | 267 (87.3) | 262 (84.8) | 254 (82.2) | 783 (84.7) |
| BMI, kg/m<sup>2</sup>, M (SD)<sup>1</sup> | n = 300; 27.66 (6.21) | n = 306; 27.27 (5.84) | n = 308; 27.25 (6.24) | n = 914; 27.39 (6.10) |
| Health food choice motives, M (SD)<sup>2</sup> | 2.74 (0.67) | 2.81 (0.70) | 2.85 (0.71) | 2.80 (0.69) |
| EDE-QS mean score, M (SD)<sup>3</sup> | 0.56 (0.57) | 0.61 (0.54) | 0.61 (0.57) | 0.60 (0.56) |
| Annual household income category score (1–8)<sup>4</sup>, M (SD) | 4.59 (1.87) | 4.72 (1.92) | 4.67 (1.94) | 4.66 (1.91) |
| Mental health diagnosis (yes), n (%) | 68 (22.2) | 74 (23.9) | 79 (25.6) | 221 (23.9) |
| **Study 2** | **WL<br>(n = 75)** | **TLL<br>(n = 82)** | **Control<br>(n = 83)** | **Overall<br>(N = 240)** |
| Age, years, M (SD) | 31.21 (9.15) | 30.27 (8.11) | 32.41 (10.72) | 31.30 (9.40) |
| Female, n (%) | 59 (78.7) | 60 (73.2) | 60 (72.3) | 179 (74.6) |
| Bachelor’s degree or above, n (%) | 55 (73.3) | 55 (67.1) | 64 (77.1) | 174 (72.5) |
| White ethnicity, n (%) | 61 (81.3) | 68 (82.9) | 70 (84.3) | 199 (82.9) |
| BMI, kg/m<sup>2</sup>, M (SD)<sup>1</sup> | n = 57; 26.63 (9.89) | n = 52; 27.16 (9.73) | n = 60; 26.96 (9.76) | n = 169; 26.92 (9.74) |
| Health food choice motives, M (SD)<sup>2</sup> | 2.83 (0.78) | 2.89 (0.83) | 2.88 (0.76) | 2.87 (0.79) |
| EDE-QS mean score, M (SD)<sup>3</sup> | 1.47 (0.61) | 1.40 (0.52) | 1.51 (0.58) | 1.46 (0.57) |
| Annual household income category score (1-8)<sup>4</sup> M (SD) | n = 73; 4.58 (1.90) | n = 80; 5.06 (1.92) | n = 77; 4.68 (2.11) | n = 230; 4.78 (1.98) |
| Mental health diagnosis, n (%) | 53 (70.7) | 61 (74.4) | 63 (75.9) | 177 (73.8) |
| Anorexia nervosa, n (%) | 31 (41.3) | 27 (32.9) | 27 (32.5) | 85 (35.4) |
| Atypical anorexia nervosa, n (%) | 5 (6.7) | 6 (7.3) | 8 (9.6) | 19 (7.9) |
| ARFID, n (%) | 7 (9.3) | 7 (8.5) | 7 (8.4) | 21 (8.8) |
| Binge-ED, n (%) | 33 (44.0) | 33 (40.2) | 32 (38.6) | 98 (40.8) |
| Bulimia nervosa, n (%) | 20 (26.7) | 23 (28.0) | 26 (31.3) | 69 (28.8) |
| Night eating syndrome, n (%) | 6 (8.0) | 8 (9.8) | 12 (14.5) | 26 (10.8) |
| Purging disorder, n (%) | 5 (6.7) | 4 (4.9) | 9 (10.8) | 18 (7.5) |
| OSFED, n (%) | 8 (10.7) | 2 (2.4) | 12 (14.5) | 22 (9.2) |
| Other ED, n (%) | 1 (1.3) | 0 (0.0) | 1 (1.2) | 2 (0.8) |

**Note.** <sup>1</sup>BMI values below 10 or above 60 kg/m<sup>2</sup> were treated as implausible and excluded from descriptive summaries. <sup>2</sup>Scored on a 4-point Likert scale (1-4), a higher score indicates greater health-related food choice motives; <sup>3</sup>Scored on a 4-point Likert scale (0-3), a higher score indicates greater ED symptomology; <sup>4</sup>1 = < £10,000, 8 = >£80,000. The category ‘£61,000 - £70,000’ was missing as a response option in error.

**Table 2:** Descriptives for primary and secondary outcomes.

| Outcome | WL | TLL | Control | Overall |
|---|---|---|---|---|
| **Study 1** |  |  |  |  |
| **Primary outcome** |  |  |  |  |
| Unhealthy food items selected, M (SD) | n = 306<br>2.34 (1.31) | n = 309<br>2.54 (1.23) | n = 309<br>2.62 (1.11) | n = 924<br>2.50 (1.22) |
| **Secondary outcomes** |  |  |  |  |
| Total calories (kcal), M (SD) | n = 306<br>1318.60 (172.49) | n = 304<br>1342.12 (166.77) | n = 308<br>1328.19 (171.46) | n = 918<br>1329.61 (170.35) |
| Total fat (g), M (SD) | n = 304<br>50.14 (10.48) | n = 303<br>51.91 (9.29) | n = 304<br>51.70 (9.26) | n = 911<br>51.25 (9.71) |
| Total saturated fat (g), M (SD) | n = 301<br>14.91 (3.92) | n = 309<br>15.62 (4.01) | n = 305<br>15.71 (3.75) | n = 915<br>15.42 (3.91) |
| Total sugar (g), M (SD) | n = 306<br>31.65 (6.64) | n = 305<br>30.52 (6.03) | n = 308<br>30.90 (6.20) | n = 919<br>31.02 (6.30) |
| Total salt (g), M (SD) | n = 306<br>4.45 (0.55) | n = 307<br>4.52 (0.53) | n = 308<br>4.50 (0.50) | n = 921<br>4.49 (0.53) |
| PANAS negative affect; pre-task, M (SD)<sup>1</sup> | n = 306<br>1.29 (0.56) | n = 309<br>1.32 (0.62) | n = 309<br>1.30 (0.56) | n = 924<br>1.30 (0.58) |
| PANAS negative affect; post-task, M (SD)<sup>1</sup> | n = 306<br>1.20 (0.47) | n = 309<br>1.25 (0.53) | n = 309<br>1.22 (0.45) | n = 924<br>1.22 (0.48) |
| Negative arousal in response to the label, M (SD)<sup>2</sup> | n = 306<br>2.97 (1.66) | n = 309<br>3.34 (1.85) | — | n = 615<br>3.16 (1.77) |
| Reactance, M (SD)<sup>2</sup> | n = 286<br>2.39 (1.62) | n = 309<br>2.29 (1.74) | — | n = 595<br>2.34 (1.69) |
| Avoidance, M (SD)<sup>2</sup> | n = 306<br>3.23 (1.90) | n = 309<br>3.33 (2.10) | — | n = 615<br>3.28 (2.00) |
| Label support, M (SD)<sup>2</sup> | n = 291<br>5.58 (1.25) | n = 303<br>5.93 (1.25) | — | n = 594<br>5.76 (1.26) |
| Perceived message effectiveness, M (SD)<sup>1</sup> | n = 296<br>2.23 (0.94) | n = 309<br>2.49 (1.14) | — | n = 605<br>2.36 (1.05) |
| **Study 2** |  |  |  |  |
| **Primary outcome** |  |  |  |  |
| Unhealthy food items selected, baseline → follow-up, M (SD) | n = 75; 2.19 (1.11) → 1.80 (1.42) | n = 82; 2.52 (1.11) → 2.39 (1.26) | n = 83; 2.40 (1.02) → 2.40 (1.10) | n = 240; 2.38 (1.09) → 2.21 (1.29) |
| **Secondary outcomes** |  |  |  |  |
| Calories, baseline → follow-up, M (SD) | n = 75; 1311.76 (171.52) → 1280.91 (159.57) | n = 82; 1306.80 (171.12) → 1311.07 (172.83) | n = 83; 1305.59 (152.15) → 1299.34 (141.55) | n = 240; 1307.93 (164.26) → 1297.59 (158.21) |
| Fat (g), baseline → follow-up, M (SD) | n = 74; 49.08 (9.24) → 44.27 (8.67) | n = 82; 49.75 (9.30) → 48.66 (9.88) | n = 83; 50.04 (7.94) → 49.47 (7.97) | n = 239; 49.64 (8.80) → 47.58 (9.12) |
| Saturated fat (g), baseline → follow-up, M (SD) | n = 66; 14.68 (3.51) → 12.39 (2.49) | n = 79; 15.32 (3.48) → 14.51 (3.46) | n = 83; 16.04 (3.74) → 15.75 (3.57) | n = 228; 15.40 (3.61) → 14.35 (3.51) |
| Sugar (g), baseline → follow-up, M (SD) | n = 68; 30.64 (5.56) → 32.02 (5.48) | n = 82; 30.91 (5.36) → 29.38 (5.90) | n = 81; 30.35 (5.55) → 30.86 (5.19) | n = 231; 30.64 (5.47) → 30.68 (5.61) |
| Salt (g), baseline → follow-up, M (SD) | n = 75; 4.46 (0.48) → 4.26 (0.61) | n = 82; 4.45 (0.52) → 4.52 (0.48) | n = 83; 4.50 (0.52) → 4.49 (0.53) | n = 240; 4.47 (0.51) → 4.43 (0.55) |
| PANAS negative affect: baseline pre-task, M (SD)<sup>1</sup> | n = 61; 2.02 (0.86) | n = 64; 1.91 (0.87) | n = 73; 2.24 (0.90) | n = 198; 2.07 (0.89) |
| PANAS negative affect: baseline post-task, M (SD)<sup>1</sup> | n = 61; 1.98 (0.82) | n = 64; 1.88 (0.83) | n = 73; 2.21 (0.98) | n = 198; 2.03 (0.89) |
| PANAS negative affect: follow-up pre-task, M (SD)<sup>1</sup> | n = 61; 1.97 (0.84) | n = 64; 1.73 (0.82) | n = 73; 1.96 (0.91) | n = 198; 1.89 (0.86) |
| PANAS negative affect: follow-up post-task, M (SD)<sup>1</sup> | n = 61; 2.09 (0.87) | n = 64; 1.80 (0.84) | n = 73; 1.99 (0.91) | n = 198; 1.96 (0.88) |
| PANAS change: baseline → follow-up task, M (SD) | n = 61; −0.05 (0.39) → 0.12 (0.28) | n = 64; −0.03 (0.52) → 0.07 (0.27) | n = 73; −0.03 (0.52) → 0.03 (0.27) | n = 198; −0.04 (0.48) → 0.07 (0.27) |
| Negative arousal in response to the label, M (SD)<sup>2</sup> | n = 75; 4.86 (1.59) | n = 82; 4.40 (1.62) | — | n = 157; 4.62 (1.62) |
| Reactance, M (SD)<sup>2</sup> | n = 59; 5.15 (1.44) | n = 82; 3.37 (2.03) | — | n = 141; 4.11 (2.01) |
| Avoidance, M (SD)<sup>2</sup> | n = 75; 4.59 (2.01) | n = 82; 3.82 (2.09) | — | n = 157; 4.18 (2.08) |
| Label support, M (SD)<sup>2</sup> | n = 75; 4.00 (2.07) | n = 78; 5.04 (1.58) | — | n = 153; 4.53 (1.91) |
| Perceived message effectiveness, M (SD)<sup>1</sup> | n = 75; 3.43 (1.18) | n = 82; 3.18 (1.15) | — | n = 157; 3.30 (1.17) |

**Note.** Label-perception outcomes were administered only in the WL and TLL conditions. Descriptive statistics are presented for the analysis sample used for each outcome following outcome-specific exclusion of outliers using the MAD criterion. Consequently, sample sizes vary slightly across outcomes. <sup>1</sup>Scored on a 5-point Likert scale (1-5), a higher score indicates greater negative affect / PME. <sup>2</sup>Scored on a 7-point Likert scale (1-7), a higher score indicates greater negative arousal / reactance / avoidance / label support.

See Appendix D for additional outcome descriptives such as label awareness, label influence, and study aim awareness.

### Primary analysis

#### Unhealthy food item selection

In Study 1 (sample without ED diagnosis), there was a significant effect of label condition on the number of task-specific unhealthy food items selected, F(2, 921) = 4.37, p = .013, η² = .009. Planned comparisons indicated that participants in the WL condition selected 0.20 fewer unhealthy items than participants in the TLL condition (p = .038, −8% difference) and 0.28 fewer than those in the control condition (p =.004, −10.7% difference). The TLL and control condition did not differ significantly (p = .428, −3% difference).

In Study 2, the condition x time interaction was not significant, F(2, 237) = 2.43, p = .090, partial η² = .020, indicating limited statistical evidence that change in unhealthy food selection differed between the three conditions. However, from baseline to follow-up, unhealthy food selection decreased by 0.39 items or 17.7% with WL (p = .015), 0.13 items or 5.3% with TLL (p =.286), and 0% with control (p= 1.00). Two-condition mixed ANOVAs indicated a greater reduction with WL than with control (mean difference in change = −0.39 items, p = .029), but not between WL and TLL (−0.25 items, p = .203) or TLL and control (−0.13 items, p = .385).

**Figure 1:**
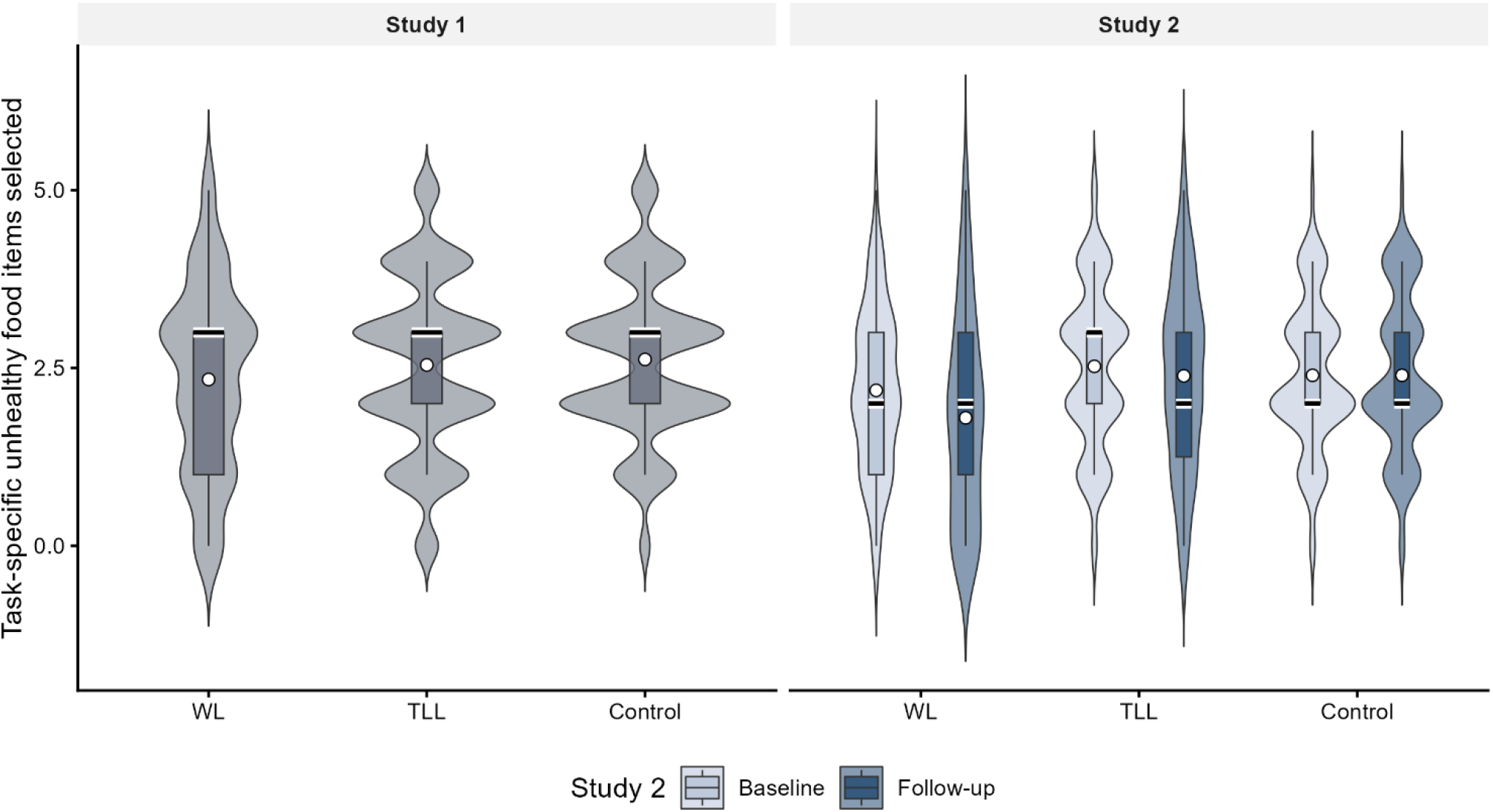
Distribution of unhealthy food items selected by label condition in Studies 1 and 2. **Note.** WL = warning label; TLL = traffic-light label. Violin plots show the distributions, boxplots show the median (black line with white outline) and interquartile range, and white points show the means.

### Secondary analysis

#### Change in negative affect

In Study 1, there was a main effect of time, F(1, 921) = 96.29, p < .001, generalized η² = .005; overall negative affect decreased from pre- food task to post-task. There was no main effect of condition, F(2, 921) = 0.46, p = .631, or condition x time interaction, F(2, 921) = 0.28, p = .757, indicating change in negative affect did not differ by label condition.

In Study 2, there was no significant condition x time x session interaction, *F*(2, 195) = 0.81, *p* = .448, partial η² = .008. This indicates that the pre-to-post-task change in negative affect between baseline and follow-up did not differ by condition.

#### Total calorie and nutrient selection

In Study 1, although individual nutrient selected tended to be lowest in the WL condition (see Figure 2), at the adjusted threshold (p <.01) label condition did not significantly affect total calories, F(2, 915) = 1.47, p = .230, η² = .003; fat, F(2, 908) = 3.05, p = .048, η² = .007; saturated fat, F(2, 912) = 3.81, p = .023, η² = .008; sugar, F(2, 916) = 2.54, p = .080, η² = .006; or salt, F(2, 918) = 1.67, p = .188, η² = .004, selected across the five trials.

**Figure 2:**
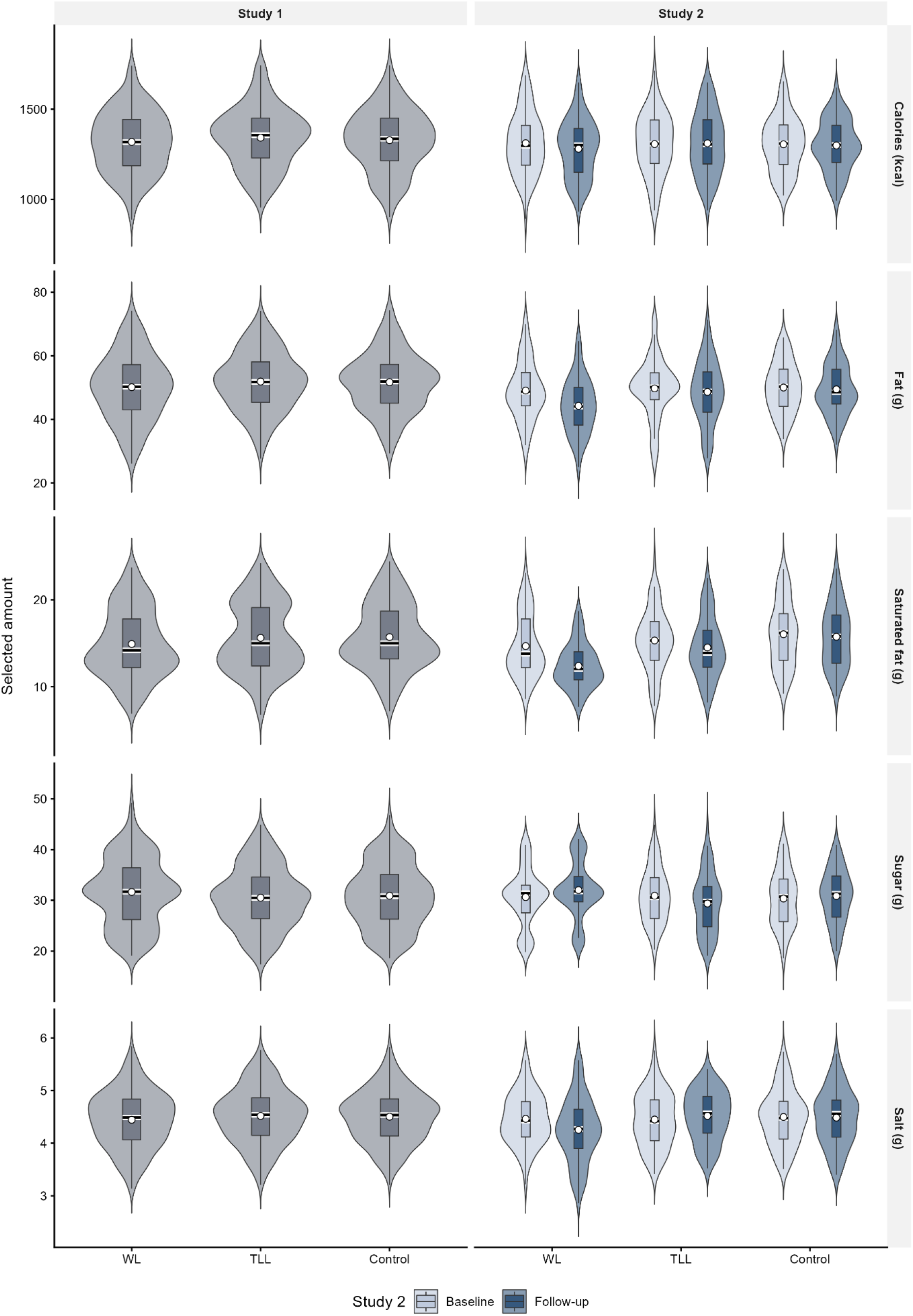
Distribution of the nutritional content of foods selected by label condition in Studies 1 and 2. **Note.** WL = warning label; TLL = traffic-light label. Violin plots show the distributions, boxplots show the median (black line with white outline) and interquartile range, and white points show the means.

In Study 2, there was no significant label condition x time interaction for total calories selected F(2, 237) = 1.00, p = .370, partial η² = .008. However, there was a significant condition x time interaction for total fat F(2, 236) = 5.54, p = .004, partial η² = .045, saturated fat *F*(2, 225) = 6.28, *p* = .002, partial η² = .053, sugar *F*(2, 228) = 4.94, *p* = .008, partial η² = .042, and salt selection *F*(2, 237) = 5.27, *p* = .006, partial η² = .043.

Planned comparisons indicated that the reduction in fat selection was greater in the WL condition than in the control condition (mean difference in change = −4.23 g, *p* = .002) and TLL condition (−3.72 g, *p* = .007). The TLL and control conditions did not differ (*p* = .704). The reduction in saturated fat selection was also greater in the WL condition than in the control condition (−2.00 g, *p* < .001). The WL versus TLL comparison did not meet the prespecified threshold (*p* = .012), and the TLL and control conditions did not differ (*p* = .340). Participants in the WL condition selected more sugar than those in the TLL condition (2.90 g, *p* = .003). Neither the WL versus control comparison (*p* = .367) nor the TLL versus control comparison (*p* = .028) were significant. Participants in the WL condition selected less salt than those in the TLL condition (−0.28 g, *p* = .002). The WL and control conditions did not differ (*p* = .025), and the TLL and control conditions did not differ (*p* = .344).

#### Perceived message effectiveness (PME)

In Study 1, PME was higher in the TLL condition than in the WL condition, F(1, 603) = 9.03, p = .003, η² = .015.

In Study 2, PME did not differ between the WL and TLL conditions, F(1, 155) = 1.78, p = .184, η² = .011.

#### Negative arousal in response to the labels

In Study 1, negative arousal was higher in the TLL condition than in the WL condition, F(1, 613) = 6.72, p = .0098, η² = .011.

In Study 2, negative arousal did not significantly differ between the WL condition and the TLL condition, *F*(1, 155) = 3.23, *p* = .074, η² = .020.

#### Reactance and avoidance

In Study 1, reactance did not significantly differ between the WL and TLL conditions, F(1, 593) = 0.56, p = .455, η² = .001. Avoidance also did not differ between the WL and TLL conditions, F(1, 613) = 0.39, p = .530, η² = .001.

In Study 2, reactance was significantly higher in the WL condition than in the TLL condition, F(1, 139) = 33.49, p < .001, η² = .194. Avoidance did not significantly differ at the prespecified statistical threshold between the WL condition and the TLL condition, *F*(1, 155) = 5.53, *p* = .020, η² = .034.

#### Acceptability of labels

In Study 1, label support was high in both label conditions but was greater in the TLL condition than in the WL condition, F(1, 592) = 11.39, p < .001, η² = .019; 79.5% of TLL participants supported the label, relative to 74.9% of WL participants.

This pattern was the same for Study 2, F(1, 151) = 12.19, p < .001, η² = .075; 66.7% of TLL participants supported the label, compared with 41.3% of WL participants.

### Sensitivity analyses

In Study 1, only 13 participants (1.4%) correctly identified the aim of the study. We reconducted the primary and secondary analyses with aim guessers removed, and findings were largely unaltered. The only change was that the negative arousal effect no longer met the adjusted threshold, F(1, 600) = 6.35, p = .012, η² = .010.

In Study 2, 35 participants (14.6%) correctly identified study aims. We reconducted the primary and secondary analyses with aim guessers removed, and the only changes were for fat, saturated fat, sugar, and salt selection, which all no longer met the adjusted threshold (p >.01). This loss of statistical significance may partly reflect reduced power following the exclusion of 35 participants, particularly for fat and sugar, for which the effect size estimates remained similar.

### Exploratory moderation analysis

Exploratory analyses examined whether age, sex, highest education (bachelor’s degree or above versus no bachelor’s degree), household income category score, and health-related food choice motives moderated the effect of label condition on unhealthy food-item selection and change in negative affect. No interaction met the preregistered p < .01 threshold in either Study 1 or Study 2 (see Appendix C).

## Discussion

Across Study 1 and 2, WL tended to outperform TLL in terms of reducing unhealthy item selection, and the amount of saturated fat and salt selected (Study 2 only). Conversely, TLL was perceived by participants to be more effective than WL for discouraging unhealthy food selection in the sample without an ED diagnosis and also preferred as a labelling format in both samples. In both studies we found no evidence that negative affect increased as a result of exposure to either labelling type. Together, findings suggest that among studied UK adults WL is most effective for changing food choice and TLL was generally preferred by participants, but neither appear to cause unintended psychological harm.

These findings are consistent with a small number of studies which indicate WL may outperform TLL in terms of reducing unhealthy food choice (3,4). We replicate these findings in a UK sample specifically, and tentatively in a UK sample with an ED diagnosis. Participants in both samples indicated greater support for TLL relative to WL as a FOP labelling policy, and this was particularly evident among participants with an ED. Participants without an ED perceived TLL to be more effective than WL, although there was no difference for participants with an ED. Theoretically it would make sense for WL to be perceived as more effective due to clearer indication of the (un)healthfulness of the product (4). It may be that because the TLL has been implemented voluntarily in the UK since 2013, UK consumers are more familiar with this label and therefore view it more favourably.

In terms of negative emotional responses to the labels, neither TLL nor WL appeared to evoke negative arousal relative to no label. When compared to each other, negative arousal was higher for TLL than WL among participants without an ED, but not among participants with an ED. Psychological reactance was higher for WL than TLL for participants with an ED, but there was no difference for participants without an ED. Other comparisons (e.g., avoidance) revealed no differences between labels or study populations. While the above outcomes produced inconsistent evidence, no change in negative affect was observed before and after being exposed to either of the labels. Collectively these findings suggest that neither acute exposure to TLL nor WL elicits notable increases in negative affect, including among individuals with an ED. This is contrary to related qualitative research which indicates that OOH calorie labelling causes emotional distress among individuals with an ED (19). It may be that because TLL and WL do not specifically focus on calories they are less distressing.

The majority female and younger age distribution of participants in Study 2 is representative of the demographics of people with an ED in the UK (17). However, this sample were also predominantly degree educated. There is limited data available on ED prevalence and education, but ED is known to impact individuals across all educational backgrounds, so this may limit generalisability. The ED sample consisted predominantly of participants reporting diagnoses of anorexia, binge eating disorder and/or bulimia. However, due to sample size we were unable to examine whether findings differed by ED type. It is therefore important to acknowledge that the impacts of labelling across ED types may not be not uniform (e.g., individuals with anorexia may be more negatively impacted by calories vs. individuals with binge eating disorder (19).

In both studies, choices were hypothetical and in the absence of price considerations, which is a limitation. However, in the real world, price would be consistent across labelling types, and hypothetical food selection task findings have been shown to directly translate into real-world behaviour (25). Moreover, in all food selection tasks except the calorie-specific task, TLL without calories were used and calorie WL were omitted. TLL were also applied to all products (under voluntary implementation coverage is approximately 75% of products (32)). This makes results less generalisable to the real-world as calorie information may or may not be required to be displayed in mandatory FOP labelling approaches. However, if this was the case it may be that both WL and TLL could evoke greater negative affect among people with an ED due to displaying calorie content (19).

From a policy perspective, our findings suggest that implementation of a mandatory WL policy in the UK could reduce selection of unhealthy food. Indeed, recent findings from Chile indicate that WL have been estimated to have reduced rates of childhood obesity in Chile since their implementation (33) and, if mandated in the UK are estimated to yield greater reductions in cardiovascular disease and obesity relative to TLL (34). WL may also be more effective than TLL for individuals with lower health literacy due to their simple, clear messaging (3). However, it would be important to engage in consultation with individuals with an ED to ascertain potential impacts of WL on ED behaviours, in addition to potential negative emotional impacts assessed here.

Implementing TLL mandatorily would be a positive (but likely less impactful on food choice) alternative step and would be received favourably by the UK public based on our findings. Due to the voluntary nature of the current TLL policy in the UK, an estimated quarter of products (often unhealthy) do not to display the label (32). Other TLL improvements indicated in related research should also be considered, including design and portion size consistency (14).

## Conclusion

Overall, WL appears to be more effective than TLL in changing hypothetical food selection among individuals without an ED, and tentatively individuals with an ED. There were no substantial negative emotional effects observed in response to either label, relative to no label, although mandatory implementation of TLL may initially be more publicly acceptable. The UK Government should consider a mandatory FOP WL strategy or substantial improvements to the current FOP TLL strategy.

## Data Availability

The study protocols and anonymised datasets are available at https://osf.io/879kh

## Contributors

RE: conceptualisation, data curation, formal analysis, investigation, methodology, project administration, validation, visualisation, writing—original draft, and writing—review and editing. RL: conceptualisation, data curation, investigation, methodology, and writing—review and editing. AF: validation and writing—review and editing. ER: conceptualisation, funding acquisition, methodology, project administration, supervision, validation, and writing—review and editing. For Study 1, RE, RL, and ER accessed and verified the data. For Study 2, RE, AF, and ER accessed and verified the data. All authors were permitted access to all the raw data in the study, and all authors accept responsibility for the decision to submit the manuscript.

## Data sharing

The study protocols and anonymised datasets are available at https://osf.io/879kh.

## Acknowledgments

This research was funded by the National Institute for Health and Care Research (NIHR) Oxford Health BRC. The views expressed are those of the author(s) and not necessarily those of the NIHR or the Department of Health and Social Care.

## Supplementary Material

### Appendix A – Prolific sample stratification plan

| Gender | Education | Age |
|---|---|---|
| Female<br>51% = 482 | NQF 2 or below<br>33% = 159 | 18 – 39<br>36% = 57<br>Ppts available = 1036<br>Cost: £119.68 |
|  |  | 40+<br>64% = 102<br>Ppts available = 1295<br>Cost: £214.18 |
|  | NQF 3<br>19% = 92 | 18 – 39<br>36% = 33<br>Ppts available = 5168<br>Cost: £69.28 |
|  |  | 40+<br>64% = 59<br>Ppts available = 2727<br>Cost: £123.88 |
|  | NQF 4+<br>48% = 231 | 18 – 39<br>36% = 83<br>Ppts available =12, 282<br>Cost: £174.28 |
|  |  | 40+<br>64% = 148<br>Ppts available = 5637<br>Cost: £310.78 |
| Male<br>49% = 464 | NQF 2 or below<br>33% = 153 | 18 – 39<br>36% = 55<br>Ppts available = 811<br>Cost: £115.48 |
|  |  | 40+<br>64% = 98<br>Ppts available = 760<br>Cost: £205.78 |
|  | NQF 3<br>19% = 88 | 18 – 39<br>36% = 32<br>Ppts available = 3648<br>Cost: £67.18 |
|  |  | 40+<br>64% = 56<br>Ppts available = 1691<br>Cost: £117.58 |
|  | NQF 4+<br>48% = 223 | 18 – 39<br>36% = 80<br>Ppts available = 7118<br>Cost: £167.98 |
|  |  | 40+<br>64% = 143<br>Ppts available = 3410<br>Cost: £300.28 |

### Appendix B – Products included in the food selection tasks

**Table B1.** Criteria for ‘high in’ nutrients.

| **Nutrient** | **High per 100g (>25% RIs)** | **High per portion (>30% of RIs)** |
|---|---|---|
| Fat | >17.5g/100g | >21g/portion |
| Saturated Fat | >5g/100g | >6.0g/portion |
| Sugar | >22.5g/100g | >27g/portion |
| Salt | >1.5g/100g | >1.8g/portion |
| Calories | >25% RI (2000 kcals) | >25% RI (2000 kcals)* |

\*For the lunch salad task, products containing 495 – 503 kcal per portion received a high calorie label due to rounding during stimulus development.

Each food choice scenario contained six products. In the crisps task, two products (33.3%) were not high in any nutrient, one (16.7%) was high in one nutrient, two (33.3%) were high in two nutrients, and one (16.7%) was high in three nutrients. In the cereal task, three products (50.0%) were not high in any nutrient and three (50.0%) were high in one nutrient. In the pasta ready-meal task, one product (16.7%) was not high in any nutrient, two (33.3%) were high in one nutrient, two (33.3%) were high in two nutrients, and one (16.7%) was high in three nutrients. In the lunch-salad task, where calories were included as the task-specific component, three products (50.0%) were high in calories only and three (50.0%) were not high in any assessed component. Finally, in the savoury-snack task, two products (33.3%) were not high in any nutrient, two (33.3%) were high in one nutrient, and two (33.3%) were high in two nutrients.

**Table B2.**
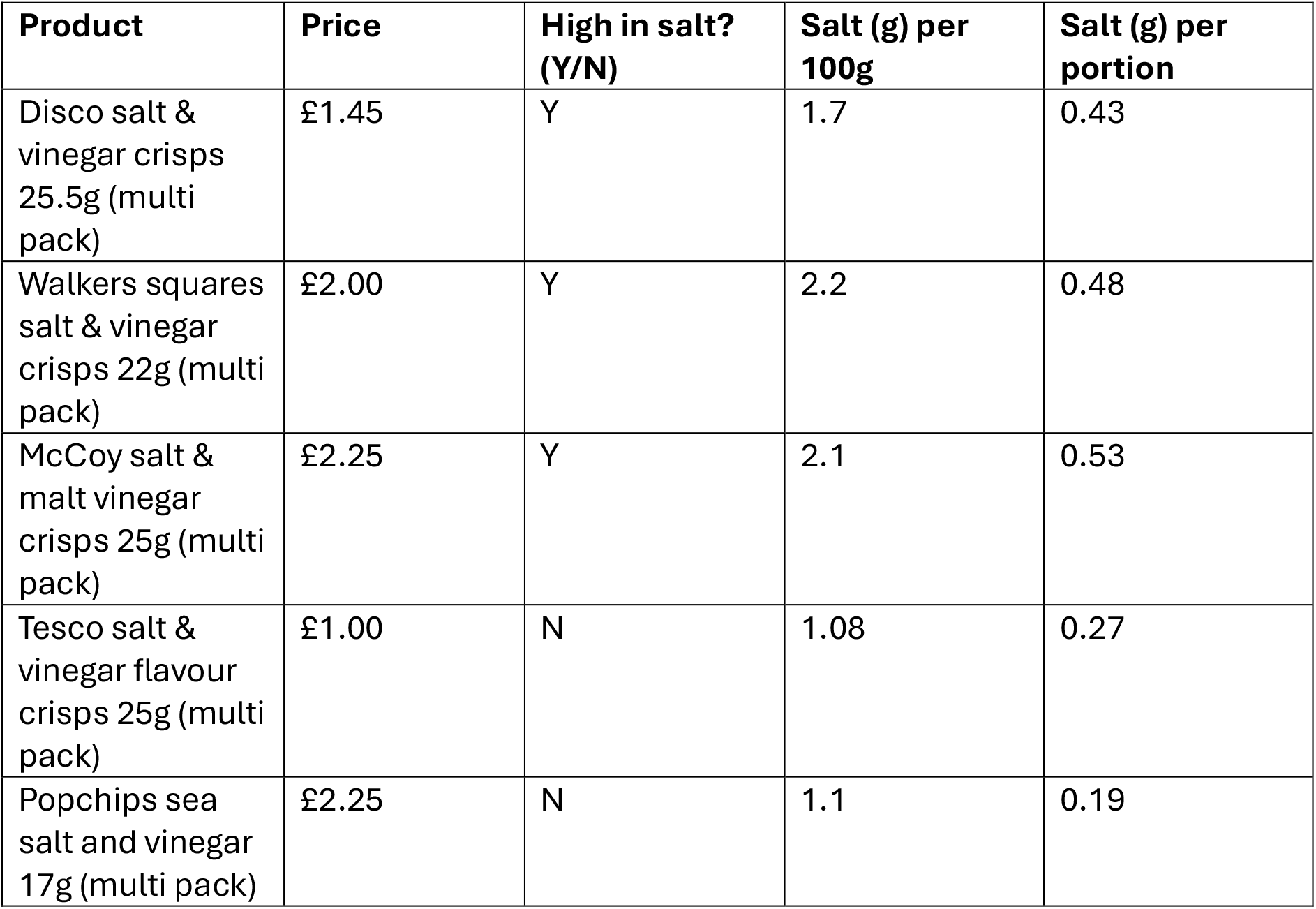
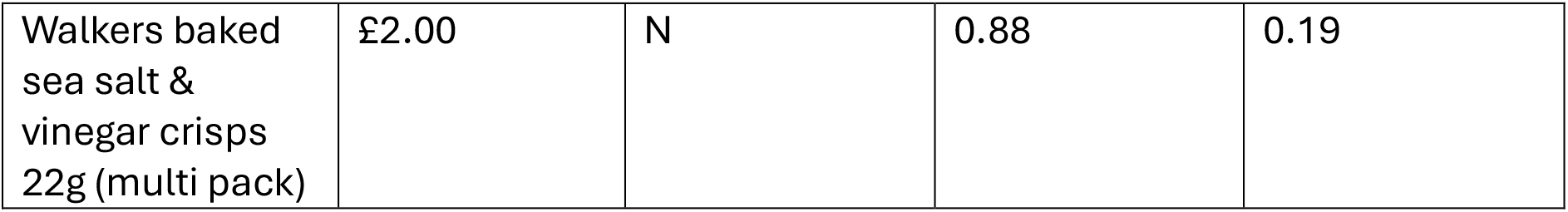
Crisps for selection.

| **Product** | **Price** | **High in salt? (Y/N)** | **Salt (g) per 100g** | **Salt (g) per portion** |
|---|---|---|---|---|
| Disco salt & vinegar crisps 25.5g (multi pack) | £1.45 | Y | 1.7 | 0.43 |
| Walkers squares salt & vinegar crisps 22g (multi pack) | £2.00 | Y | 2.2 | 0.48 |
| McCoy salt & malt vinegar crisps 25g (multi pack) | £2.25 | Y | 2.1 | 0.53 |
| Tesco salt & vinegar flavour crisps 25g (multi pack) | £1.00 | N | 1.08 | 0.27 |
| Popchips sea salt and vinegar 17g (multi pack) | £2.25 | N | 1.1 | 0.19 |
| Walkers baked sea salt & vinegar crisps 22g (multi pack) | £2.00 | N | 0.88 | 0.19 |

**Figure B1.**
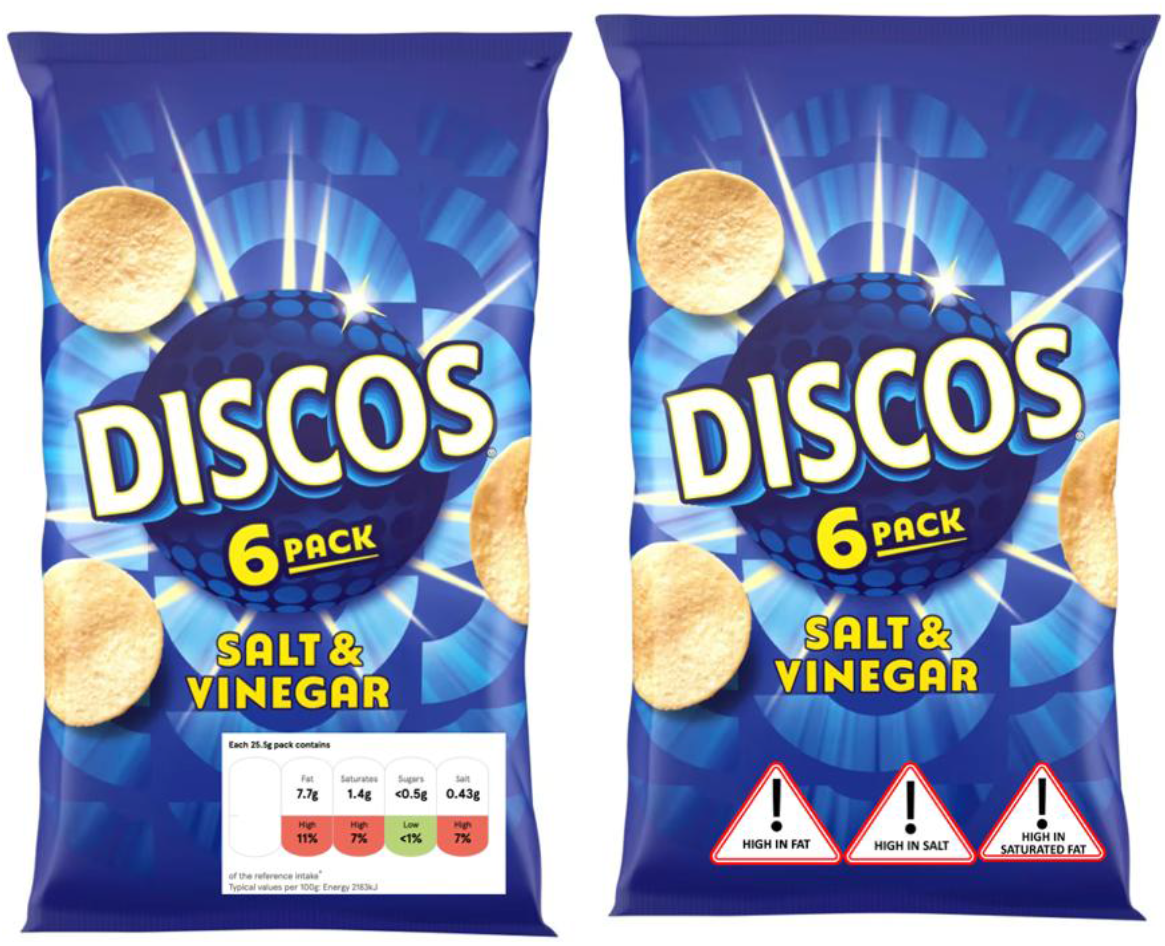
Example of crisps for selection

**Table B3.**
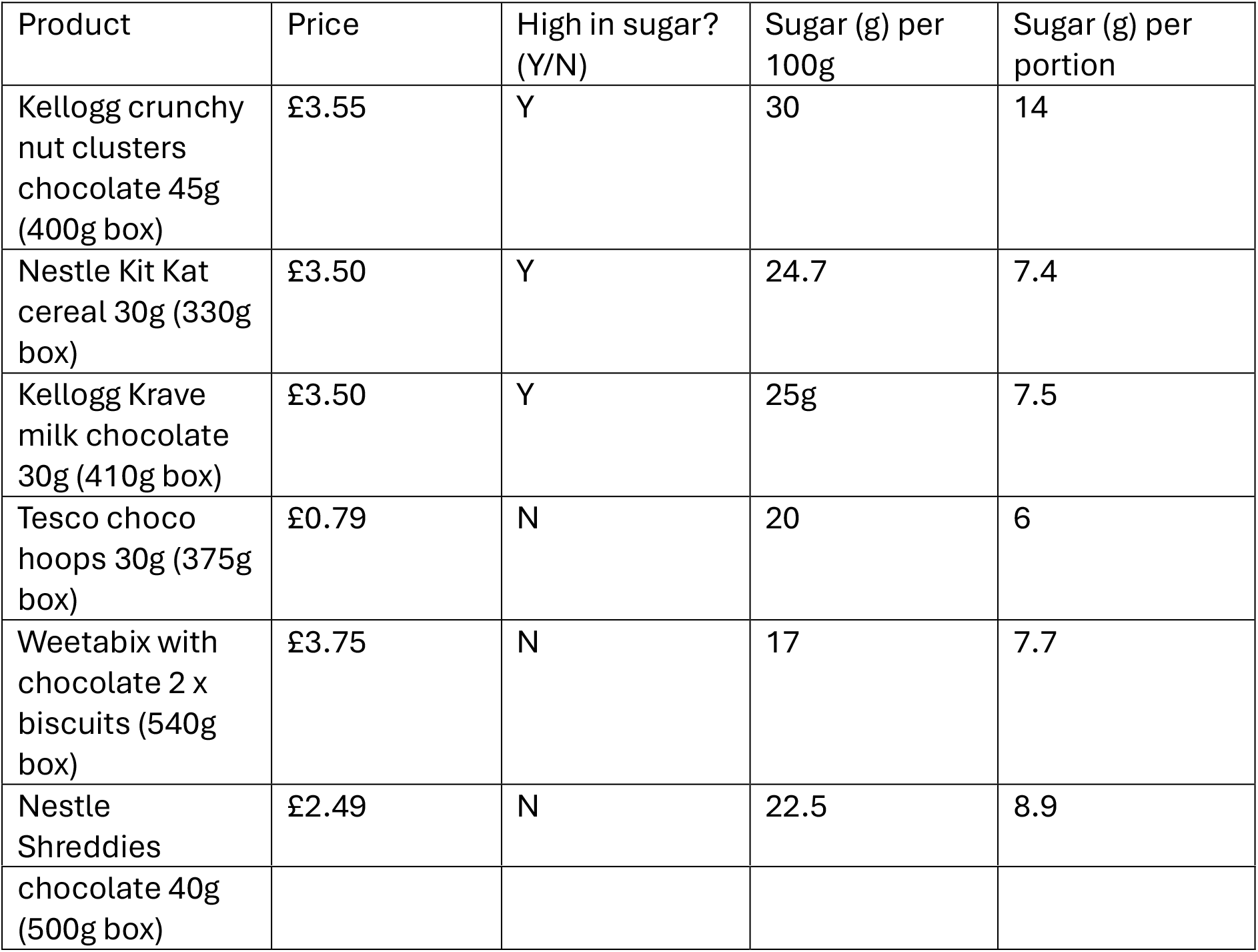
Cereals for selection.

| Product | Price | High in sugar? (Y/N) | Sugar (g) per 100g | Sugar (g) per portion |
|---|---|---|---|---|
| Kellogg crunchy nut clusters chocolate 45g (400g box) | £3.55 | Y | 30 | 14 |
| Nestle Kit Kat cereal 30g (330g box) | £3.50 | Y | 24.7 | 7.4 |
| Kellogg Krave milk chocolate 30g (410g box) | £3.50 | Y | 25g | 7.5 |
| Tesco choco hoops 30g (375g box) | £0.79 | N | 20 | 6 |
| Weetabix with chocolate 2 x biscuits (540g box) | £3.75 | N | 17 | 7.7 |
| Nestle Shreddies | £2.49 | N | 22.5 | 8.9 |
| chocolate 40g (500g box) |  |  |  |  |

**Figure B2.**
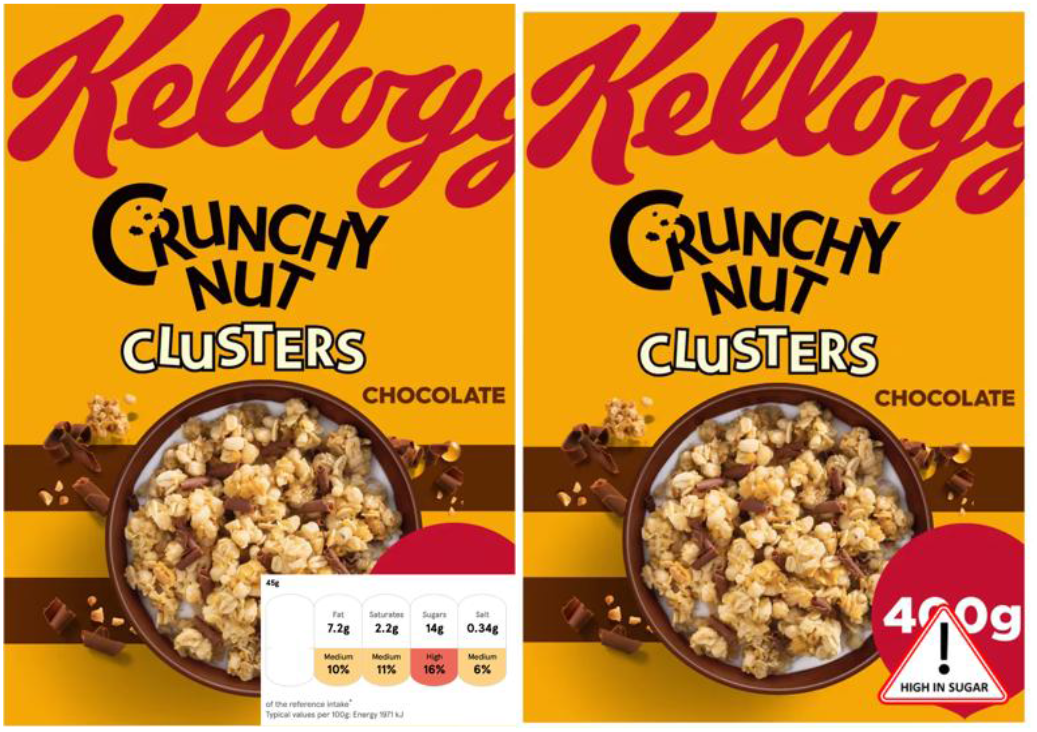
Example of cereal for selection

**Table B4.** Savoury snacks for selection.

| Product | Price | High in fat? (Y/N) | Fat (g) per 100g | Fat (g) per portion |
|---|---|---|---|---|
| Tesco Chargrilled artichokes ¼ pack of 140g | £3.00 | Y | 22.6 | 7.9 |
| Tesco mozzarella & slow roasted tomatoes 1/3 pack of 200g | £3.00 | Y | 20 | 13.4 |
| Tesco Italian nocellara olives ¼ 140g pot | £3.00 | Y | 19 | 6.7 |
| Tesco Spanish style tortilla ¼ of 500g pack | £3.00 | N | 8.8 | 11 |
| Tesco stuffed vine leaves 1 in 162g pot | £3.00 | N | 4.9 | 1.3 |
| Tesco soft cheese stuffed peppers ¼ of 145g pack | £3.00 | N | 14.6 | 5.2 |

**Figure B3.**
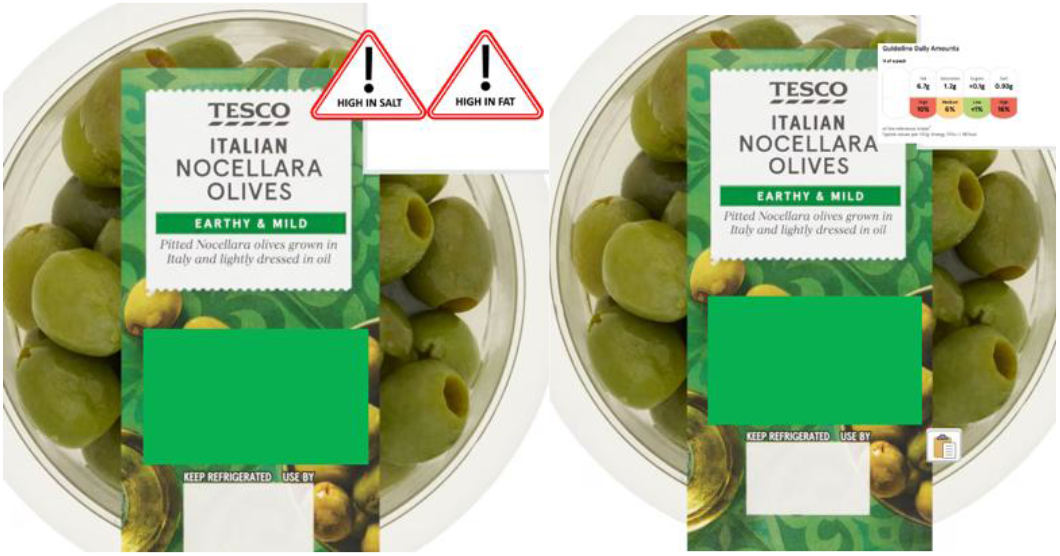
Example of savoury snack for selection

**Table B5.** Pasta ready meals for selection.

| Product | Price | High in saturated fat? (Y/N) | Saturated fat (g) per 100g | Saturated fat (g) per portion |
|---|---|---|---|---|
| Tesco spinach & ricotta cannelloni 400g | £3.30 | Y | 2.8 | 11.0 |
| Tesco spaghetti carbonara 400g | £3.30 | Y | 1.7 | 6.8 |
| Tesco beef lasagne 400g | £3.30 | Y | 3.2 | 12.9 |
| Tesco pepperoni pasta bake | £3.30 | N | 1.4 | 5.7 |
| Tesco ham & mushroom tagliatelle 400g | £3.30 | N | 1.3 | 5.2 |
| Tesco tomato & mozzarella pasta bake 400g | £3.30 | N | 1.3 | 5.3 |

**Figure B4.**
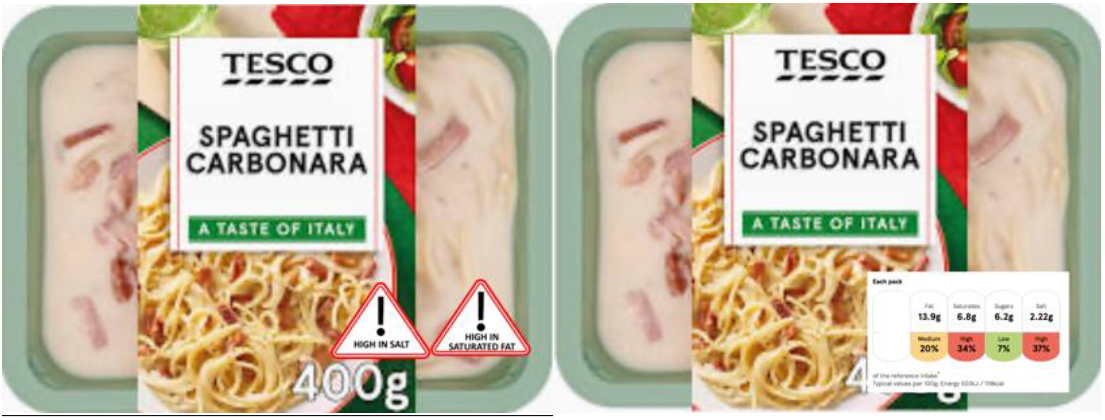
Example of pasta ready meal for selection

**Table B6.** Lunch salad for selection.

| Product | Price | High in calories? (Y/N) | Calories (g) per 100g | Calories (g) per portion |
|---|---|---|---|---|
| Tesco feta & semi dried tomato pasta 290g | £2.85 | Y | 171 | 495 |
| Tesco southern fried chicken pasta 300g | £2.85 | Y | 166 | 498 |
| Tesco chicken and smoked bacon pasta 300g | £2.85 | Y | 168 | 503 |
| Tesco hoisin duck noodle bowl 250g | £3.00 | N | 72 | 179 |
| Tesco fajita fiesta chicken salad 265g | £3.00 | N | 126 | 333 |
| Tesco eat your greens feta salad 240g | £3.60 | N | 126 | 302 |

**Figure B7.**
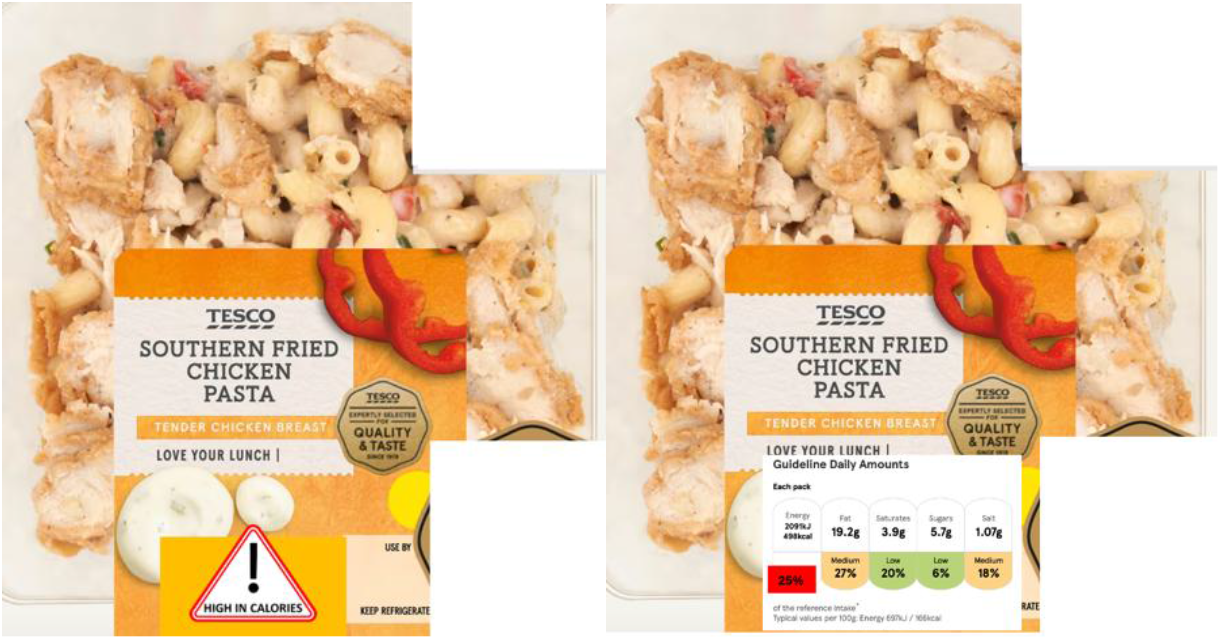
Example of Lunch salad for selection

### Appendix C - Moderation analysis detail

**Table C1.** Moderation analysis.

| **Outcome** | **Moderator** | **Study 1 interaction test** | **Study 2 interaction test** |
|---|---|---|---|
| **Number of unhealthy food items selected** | Age | F(2, 918) = 0.32, p = .724 | F(2, 233) = 0.94, p = .391 |
|  | Sex | F(2, 916) = 4.06, p = .018 | F(2, 233) = 1.92, p = .149 |
|  | Highest education | F(2, 918) = 3.26, p = .039 | F(2, 233) = 0.93, p = .397 |
|  | Household income | F(2, 918) = 0.81, p = .446 | F(2, 223) = 1.25, p = .288 |
|  | Health food choice motives | F(2, 918) = 4.07, p =.017 | F(2, 233) = 1.02, p = .364 |
| **Change in negative affect** | Age | F(2, 918) = 3.32, p = .037 | F(2, 191) = 0.07, p = .934 |
|  | Sex | F(2, 916) = 2.62, p = .073 | F(2, 191) = 0.11, p = .897 |
|  | Highest education | F(2, 918) = 0.45, p = .637 | F(2, 191) = 0.71, p = .495 |
|  | Household income | F(2, 918) = 0.83, p = .434 | F(2, 181) = 0.25, p = .776 |
|  | Health-related food choice motives | F(2, 918) = 0.44, p = .643 | F(2, 191) = 0.40, p = .673 |

### Appendix D – Additional Study 1 and Study 2 outcomes

**Table D1.**
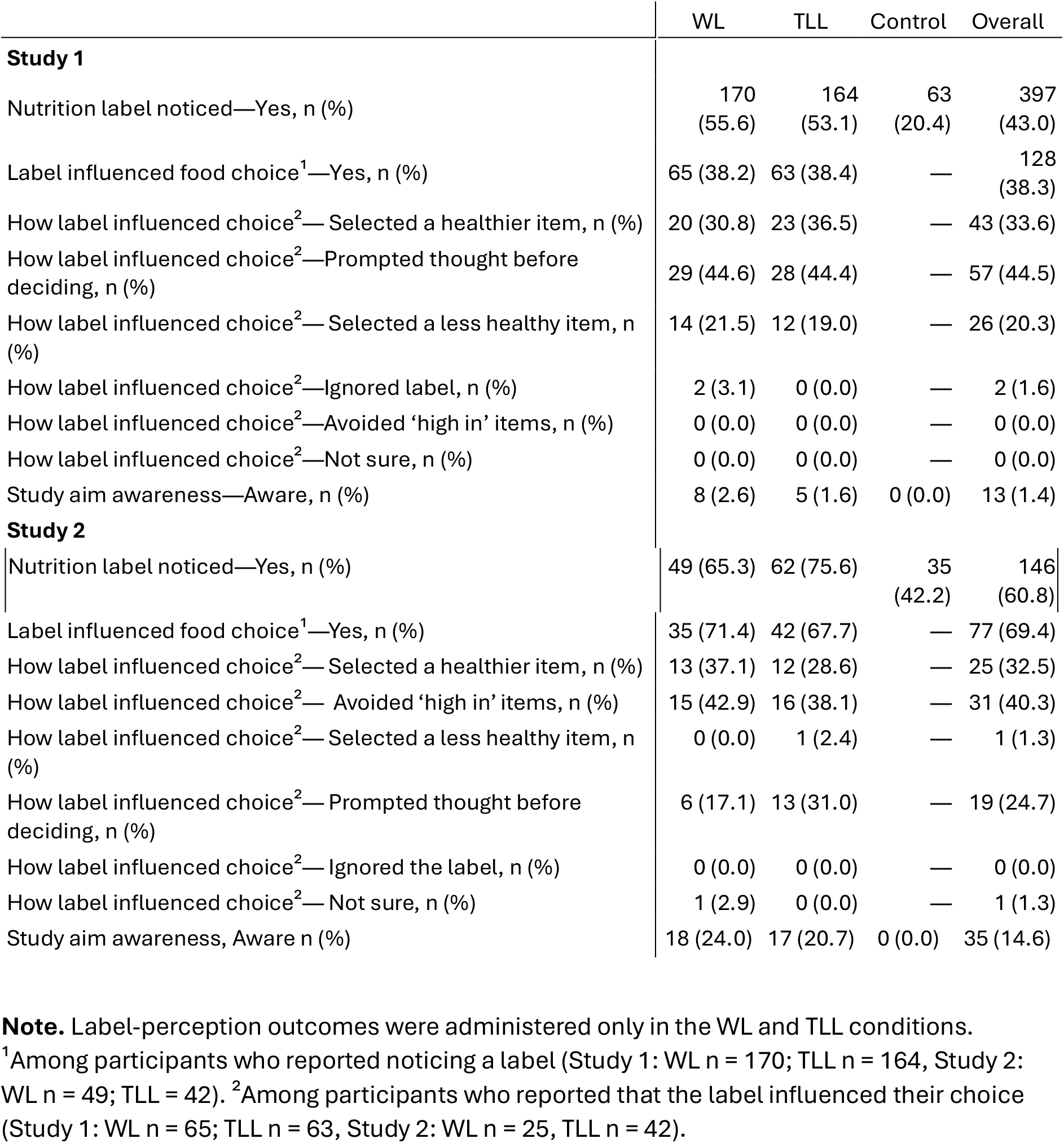
Descriptives for additional Study 1 and Study 2 outcomes.

### Appendix E – Sensitivity ANCOVAs for Study 2

#### Unhealthy food item selection

In Study 2, an ANCOVA controlling for the number of unhealthy food items selected during the baseline no-label task indicated a significant effect of follow-up label condition, F(2, 236) = 4.12, p = .017, partial η² = .034. Participants in the WL condition selected fewer unhealthy food items at follow-up (M = 1.80, SD = 1.42) than participants in the TLL (M = 2.39, SD = 1.26) and control conditions (M = 2.40, SD = 1.10). Planned comparisons of baseline-adjusted means indicated that participants in the WL condition selected fewer unhealthy items than participants in the control condition (adjusted mean difference = −0.46, p = .007) and TLL condition (adjusted mean difference = −0.37, p = .030). The TLL and control condition did not significantly differ (adjusted mean difference = −0.09, p = .581).

#### Total calorie and nutrient selection

In Study 2, there was also no significant effect of label condition on total calories selected *F*(2, 236) = 1.16, p = .316, partial η² = .010. However, there was a significant effect of label condition on total fat (*F*(2, 235) = 8.65, *p* < .001, partial η² = .069), saturated fat (F(2, 224) = 17.11, p < .001, partial η² = .133), sugar (F(2, 227) = 5.86, p = .003, partial η² = .049), and salt selection (F(2, 236) = 6.81, p = .001, partial η² = .055).

Planned comparisons indicated that participants in the WL condition selected less fat than participants in the TLL condition (adjusted mean difference = −4.03 g, p = .001) and control condition (adjusted mean difference = −4.68 g, p < .001). TLL and control participants did not differ (p = .583). Participants in the WL condition selected less saturated fat than those in the TLL condition (adjusted mean difference = −1.84 g, p < .001) and control condition (adjusted mean difference = −2.77 g, p < .001). The TLL and control conditions did not differ at the prespecified threshold (p = .040). Participants in the WL condition selected more sugar than participants in the TLL condition (adjusted mean difference = 2.75 g, p <.001). Neither the WL versus control comparison (p = .209) nor the TLL versus control comparison (p = .030) was significant at the prespecified threshold. Participants in the WL condition selected less salt than those in the TLL (adjusted mean difference = −0.27 g, p = <.001) and control condition (adjusted mean difference = −0.22 g, p = .006), The TLL and control conditions did not differ (p = .457).

#### Change in negative affect

In Study 2, an ANCOVA controlling for change in negative affect during the baseline no-label task found no evidence that follow-up change in negative affect differed between conditions, F(2, 194) = 2.04, p = .132, partial η² = .021.

## References

1. Department of Health and Social Care. Public Health Profiles - Obesity, Physical Activity, and Nutrition. 2026. doi:https://fingertips.phe.org.uk/profile/obesity-physical-activity-nutrition

2. House of Lords Food, Diet and Obesity Committee. Recipe for health: a plan to fix our broken food system [Internet]. 2024 Oct. Available from: https://committees.parliament.uk/committee/698/food-diet-and-obesity-committee/news/203372/the-government-needs-a-plan-to-fix-our-broken-food-system-and-turn-the-tide-on-the-public-health-emergency/

3. Croker H, Packer J, Russell SJ, Stansfield C, Viner RM. Front of pack nutritional labelling schemes: a systematic review and meta-analysis of recent evidence relating to objectively measured consumption and purchasing. J Hum Nutr Diet. 2020 Aug;33(4):518–37. doi:10.1111/jhn.12758

4. Song J, Brown MK, Tan M, MacGregor GA, Webster J, Campbell NRC, et al. Impact of color-coded and warning nutrition labelling schemes: A systematic review and network meta-analysis. PLOS Med. 2021;18(10):e1003765. doi:10.1371/journal.pmed.1003765

5. Roberto CA, Ng SW, Ganderats-Fuentes M, Hammond D, Barquera S, Jauregui A, et al. The Influence of Front-of-Package Nutrition Labeling on Consumer Behavior and Product Reformulation. Annu Rev Nutr. 2021 Oct 11;41(1):529–50. doi:10.1146/annurev-nutr-111120-094932

6. Food Standards Agency. Check the label. How to use nutritional labels on pre-packed foods to find calorie, fat, saturates, sugars and salt content information. [Internet]. 2020. Available from: https://www.food.gov.uk/safety-hygiene/check-the-label

7. Fichera E, Von Hinke S. The response to nutritional labels: Evidence from a quasi-experiment. J Health Econ. 2020 Jul;72:102326. doi:10.1016/j.jhealeco.2020.102326

8. Reyes M, Garmendia ML, Olivares S, Aqueveque C, Zacarías I, Corvalán C. Development of the Chilean front-of-package food warning label. BMC Public Health. 2019;19(1):1–11.

9. GOV.UK. Front-of-pack nutrition labelling in the UK: building on success [Internet]. 2020. Available from: https://www.gov.uk/government/consultations/front-of-pack-nutrition-labelling-in-the-uk-building-on-success

10. Taillie LS, Bercholz M, Popkin B, Rebolledo N, Reyes M, Corvalán MC. Decreases in purchases of energy, sodium, sugar, and saturated fat 3 years after implementation of the Chilean food labeling and marketing law: An interruptd time series analysis. PLOS Med. 2024 Sep 27;21(9):e1004463. 10.1371/journal.pmed.1004463

11. Salgado JC, Pedraza LS, Contreras-Manzano A, Aburto TC, Tolentino-Mayo L, Barquera S. Product reformulation in non-alcoholic beverages and foods after the implementation of front-of-pack warning labels in Mexico. PLoS Med. 2025;22(3):e1004533.

12. Saavedra-Garcia L, Meza-Hernández M, Diez-Canseco F, Taillie LS. Reformulation of Top-Selling Processed and Ultra-Processed Foods and Beverages in the Peruvian Food Supply after Front-of-Package Warning Label Policy. Int J Environ Res Public Health. 2022;20(1):424.

13. Khandpur N, Mais LA, de Morais Sato P, Martins APB, Spinillo CG, Rojas CFU, et al. Choosing a front-of-package warning label for Brazil: a randomized, controlled comparison of three different label designs. Food Res Int. 2019;121:854–61.

14. Strange A, Davies S, Bellini P. Making front of pack nutrition labelling work for consumers - Which? 2026. doi:https://www.which.co.uk/policy-and-insight/article/making-front-of-pack-nutrition-labelling-work-for-consumers-ajANc1R2eh2q#chapter-1-status-quo-how-consumers-respond-to-mtl-labels

15. Sanjari SS, Jahn S, Boztug Y. Dual-process theory and consumer response to front-of-package nutrition label formats. Nutr Rev. 2017;75(11):871–82.

16. Clarke N, Pechey E, Kosīte D, König LM, Mantzari E, Blackwell AKM, et al. Impact of health warning labels on selection and consumption of food and alcohol products: systematic review with meta-analysis. Health Psychol Rev. 2021 Jul 3;15(3):430–53. doi:10.1080/17437199.2020.1780147

17. Beat. HOW WE ESTIMATED HOW MANY PEOPLE IN THE UK HAVE AN EATING DISORDER. 2026. doi:https://www.beateatingdisorders.org.uk/get-information-and-support/about-eating-disorders/how-many-people-eating-disorder-uk/eating-disorder-statistics/

18. Trompeter N, Duffy F, Peebles I, Wadhera E, Chambers K, Sharpe H, et al. Impact of out-of-home nutrition labelling on people with eating disorders: a systematic review and meta-synthesis. BMJ Public Health. 2025 Jan;3(1):e000862. doi:10.1136/bmjph-2023-000862

19. Trompeter N, Nadarajah A, Sharpe H, Duffy F, Maloney E, Nicholls D, et al. Views of people with disordered eating on current and alternative out-of-home calorie labelling policies in England: a mixed-methods survey. BMJ Public Health. 2026 Apr;4(2):e003666. doi:10.1136/bmjph-2025-003666

20. Penzavecchia C, Todisco P, Muzzioli L, Poli A, Marangoni F, Poggiogalle E, et al. The influence of front-of-pack nutritional labels on eating and purchasing behaviors: a narrative review of the literature. Eat Weight Disord - Stud Anorex Bulim Obes. 2022 Nov 11;27(8):3037–51. doi:10.1007/s40519-022-01507-2

21. ONS. Age groups. 2023.

22. ONS. Male and female populations. 2023.

23. ONS. Education and training statistics for the UK. 2023.

24. FSA. Guide to creating a front of pack (FoP) nutrition label for pre-packed products sold through retail outlets. 2016.

25. Evans R, Brealey J, Clarke N, Falbe J, Finlay A, Jones A, et al. Salt warning labels in the out-of-home food sector: online and real-world randomised controlled trials in the UK. Lancet Public Health. 2025 Aug;10(8):e656–67. doi:10.1016/S2468-2667(25)00143-4

26. Steptoe A, Pollard TM, Wardle J. Development of a Measure of the Motives Underlying the Selection of Food: the Food Choice Questionnaire. Appetite. 1995 Dec;25(3):267–84. doi:10.1006/appe.1995.0061

27. Gideon N, Hawkes N, Mond J, Saunders R, Tchanturia K, Serpell L. Development and Psychometric Validation of the EDE-QS, a 12 Item Short Form of the Eating Disorder Examination Questionnaire (EDE-Q). Takei N, editor. PLOS ONE. 2016 May 3;11(5):e0152744. doi:10.1371/journal.pone.0152744

28. Clarke N, Pechey E, Mantzari E, Blackwell AKM, De-loyde K, Morris RW, et al. Impact of health warning labels on snack selection: An online experimental study. Appetite. 2020 Nov;154:104744. doi:10.1016/j.appet.2020.104744

29. Mackinnon A, Jorm AF, Christensen H, Korten AE, Jacomb PA, Rodgers B. A short form of the Positive and Negative Affect Schedule: evaluation of factorial validity and invariance across demographic variables in a community sample. Personal Individ Differ. 1999 Sep;27(3):405–16. doi:10.1016/S0191-8869(98)00251-7

30. Baig SA, Noar SM, Gottfredson NC, Boynton MH, Ribisl KM, Brewer NT. UNC Perceived Message Effectiveness: Validation of a Brief Scale. Ann Behav Med. 2019 Jul 17;53(8):732–42. doi:10.1093/abm/kay080

31. Leys C, Ley C, Klein O, Bernard P, Licata L. Detecting outliers: Do not use standard deviation around the mean, use absolute deviation around the median. J Exp Soc Psychol. 2013 Jul;49(4):764–6. doi:10.1016/j.jesp.2013.03.013

32. FSA. Consumer responses to food labelling: A rapid evidence review. 2021.

33. Paraje G, Valdés N, Macaya AV, Corvalán C, Popkin B. The impact of Chile’s multipronged food labelling and advertising law on early childhood excess weight: a cohort difference-in-differences study. The Lancet. 2026 Jun;407(10548):2630–40. doi:10.1016/S0140-6736(26)00651-3

34. Evans R, O’Flaherty M, Putra IGNE, Kypridemos C, Robinson E, Colombet Z. The estimated impact of mandatory front-of-pack nutrition labelling policies on adult obesity prevalence and obesity-related mortality in England: a modelling study. Lancet Reg Health - Eur. 2026 Jan;60:101506. doi:10.1016/j.lanepe.2025.101506

